# Patients at risk of pulmonary fibrosis Post Covid-19: Epidemiology, pulmonary sequelaes and humoral response

**DOI:** 10.1101/2022.03.04.22271920

**Authors:** Miriam Hernández Porto, Teresa Delgado, Armando Aguirre-Jaime, María Jose Ramos, Silvia Campos, Orlando Acosta, Ana Belén Llanos, María Lecuona

## Abstract

**Background:** The COVID-19 pandemic is one of the greatest public health problems. Our aims were to describe epidemiological characteristics, know the amount of protective antibodies and their permanence after a COVID-19 primary infection in patients with risk of pulmonary fibrosis.

**Methods:** Descriptive epidemiological and follow-up study of the humoral response in patients at risk of pulmonary fibrosis Post-Covid-19 hospitalized, between March and October 2020, and who were followed up for a one year after hospital discharge.

**Results:** 72 patients participated in the study, 52 showed pre-existing chronic comorbidities. COVID-19 clinical severity was rated in 6% mild, 58% as moderate and 36% as severe. After a year of follow-up, the forty percent had pulmonary sequelae, the most frequent (20%) was mild pulmonary fibrosis. Any case of reinfection was detected. All patients presented RBD IgG antibodies and 88% presented IgA antibodies after 8-9 months. The amount of RBD IgG was similar at 4-5 and 8-9 months post-Covid. There was no difference when level of RBD IgG according to the severity of the COVID-19 (p=0.441, p=0.594).

**Conclusions:** Mild pulmonary fibrosis sequelae is exceptional but was detected in a high percentage. The amount of RBD IgG is maintained throughout the convalescent phase and seems to protect against new reinfections despite of emerging viral variants. However, seems not predict the developed or not of pulmonary fibrosis.

## Introduction

The COVID-19 pandemic continues to spread worldwide and is one of the greatest public health problems in the world. The severity of the COVID-19 picture is probably due to a previous deterioration of the immune system [1] due to the comorbidities, such as those reported Williamson et al., [2]: cardiovascular disease, diabetes, respiratory disease including severe asthma, obesity, history of hematological malignancy, cancer, kidney, liver, neurological and autoimmune conditions. Many patients are symptomatic to some degree after COVID-19 Infection but Pulmonary fibrosis is exceptional [3].

Animal models have demonstrated how SARS CoV-2 infection in macaques [4] provides effective protection against reinfection by this virus, probably due to the rapid immune control that occurs thanks to the neutralizing action of the antibodies. This action is centered on its ability to neutralize the receptor-binding domain (RBD) of the virus in the S1 subunit, thus preventing its binding to the cellular receptor angiotensin-converting enzyme 2 (ACE2) and therefore avoiding the entry of the virus into the cell. In addition, several studies have been carried out on the early use of plasma from convalescent patients with high titers of neutralizing antibodies as a therapeutic option in patients with COVID-19 [5,6].

Finally, the risk of reinfection is considerable, due to several reasons: the absence of a threshold of antibodies that predict protection, the permanence of these antibodies in the organism after a primary infection and the emerging viral variants.

Our aims were to describe epidemiological characteristics, know the amount of protective antibodies and their permanence after COVID-19 primary infection in patients with risk of pulmonary fibrosis.

## Materials and Methods

Descriptive characteristics and follow-up results of the humoral response in patients with a diagnosis of SARS-CoV-2 confirmed by RT-PCR and hospitalized at the Hospital Universitario de Canarias (Spain), between March and October 2020, and who were followed up after hospital discharge at the Multidisciplinary Interstitial Lung Disease Unit of this Hospital. These patients had to present at least one of the following conditions to be referred to such consultation: persistence of pathological alterations in the chest X-ray and/or having required special ventilatory support devices during their admission (high-flow nasal spectacles, noninvasive ventilation or intubation and mechanical ventilation). This follow-up ended when respiratory clinical normalization was observed and complete or almost complete involution of the radiological alterations initially visualized was confirmed. During follow-up and for the evaluation of possible post COVID-19 pulmonary sequelae pulmonary function tests and imaging tests (pulmonary ultrasound and high resolution computed axial tomography) were performed at 6 weeks (in all cases) and at 3-6 months and one year (those who had to be followed up due to incomplete recovery in the initial and successive visits).

Patient volunteers signed their informed consent and were subsequently scheduled for serum sampling at 4-5 months and 8-9 months after COVID-19 infection.

We analyzed different antibodies; RBD-specific IgG, Nucleocapsid IgG, and Spike 1-RBD IgM antibodies determined by Abbott chemiluminescent microparticle assays (CMIA): SARS-CoV-2 IgG II Quant, SARS-CoV-2 IgG and SARS-CoV-2 IgM using the ARCHITECT i 2000 SR system, following the manufacturer’s instructions.

IgG RBD measurements were transformed to the WHO [7] international standard BAU/mL in order to obtain comparable of antiSARS-CoV-2 antibody quantification at the international level.

IgA Spike and IgM Nucleocapsid antibodies were determined using EUROIMMUN enzyme-linked immunosorbent assay (ELISA); Anti-SARS-CoV-2 ELISA IgA and Anti-SARS-CoV-2 NCP ELISA IgM (Euroinmmun, Lübeck, Germany) according to the manufacturer’s instructions on the Dynex DS2 ELISA System platform.

Clinical variables of the patients were also collected by reviewing the medical history such as: sex and age, comorbidities considered as risk factors for worse prognosis [2], being a smoker or former smoker, degree of severity of COVID-19 disease according to the WHO guidelines (Clinical Management of COVID-19 Patients-Interim Guidance)[8], hospital admission service, hospital stay, duration of the acute phase of the disease, development of pneumonia, measurement of oxygen saturation and type of ventilatory support required during admission, months of post-discharge follow-up, clinical resolution after one year of follow-up, reinfection with SARS-CoV-2.

The characteristics of the sample are presented by summarizing the nominal variables with the absolute and relative frequency of their component categories and the numerical scale variables with mean(SD) or median(minimum-maximum) according to their normal or non-normal distribution confirmed with the Kolmogorov-Smirnov test. Comparisons of the changes in the frequencies of the ranges of antibody determinations according to each specific cut-off point from 4-5 to 8-9 months were performed with the Pearson’s chi-squared test or Fisher’s exact test. Comparisons of changes in IgG measured in BAU/ml compared to RBD from 4-5 to 8-9 months, in general, and stratified by COVID-19 severity were performed with the Wilcoxon test for paired samples. Comparison of these same determinations for the same period according to COVID-19 severity was performed with the U Mann-Whitney test.

All hypothesis contrast tests were bilateral at a significance level p≤0.05 and the calculations involved were performed with the support of the SPSS 25.0™ statistical data processing package from IBM Co®.

### Ethical Approval statement

The study was approved by Ethical Committee with the code CHUC_2020_68.

## Results

A total of 72 patients who met the inclusion criteria and gave their consent participated in the study. The total sample of participants had an age of 60(30) years in a range of 32-89 years, 53% were women. Seventy nine percent of the patients showed pre-existing chronic comorbidities. The distribution of comorbidities was as follows: 58% hypertension, 39% type 2 diabetes, 19% chronic pulmonary disease, 15% heart disease, 10% chronic kidney disease, 7% oncologic disease and 1% cerebrovascular disease. A total of 75% had 2 or more comorbidities. Some 36% were smokers or former smokers. The degree of clinical severity of COVID-19 was rated as mild in 6% mild, moderate in 58% and severe in 36%. Ninety four percent of the patients with pre-existing chronic comorbidities, showed moderate and severe COVID-19.

Patients were hospitalized in different departments: 54% in Internal Medicine-Infectious Diseases, 25% in Pneumology, 18% in Intensive Care Units and 3% in Home Hospitalization Unit. Hospital stay was 22 (1-41) days. The duration of the acute phase was 25(8-62) days. During the acute phase of the disease, 94% developed pneumonia. Forty-nine percent required no ventilatory support, while 25% required high-flow nasal spectacles, 13% noninvasive mechanical ventilation and 13% invasive according to the severity of the process. In cases with moderate and severe COVID the oxygen saturation during admission was 88(0,7) % in a range of 70-98%. 13 % of patients are still under clinical follow-up due to post COVID-19 pulmonary sequelae after one year and in the rest of patients the post-discharge clinical follow-up time by the Multidisciplinary Unit of Interstitial Lung Diseases of the Hospital was 9(1-12) months.

Of total patients, forty percent had pulmonary sequelae, including seventeen percent with minimal parenchymal alterations with no clinical or functional repercussions, three percent with had ground-glass infiltrates and twenty percent mild with pulmonary fibrosis.

40 out of 72 patients participated in the antibody analysis; 20 attended the sample collection for antibody determination at 4-5 months and at 8-9 months post COVID-19 and other 20 at 8-9 months post COVID-19. All of them had moderate or severe of COVID, and thirty six percent had minimal parenchymal sequelae with no clinical or functional repercussions, and no one developed pulmonary fibrosis.

The results of the humoral response to SARS-CoV-2 at 4-5 months and 8-9 months post Covid-19 infection are shown in Table 1.

**Table 1.** Humoral response to SARS-CoV-2 at 4-5 months and 8-9 months after Covid-19 infection.

| DETERMINATION<br>n(%) | At 4-5 months post infection |  |  |  | At 8-9 months post infection |  |  |  |
| --- | --- | --- | --- | --- | --- | --- | --- | --- |
|  | POSITIVE | NEGATIVE | UNDET. | INDEX<br>median<br>(min-max) | POSITIVE | NEGATIVE | UNDET. | INDEX<br>median<br>(min-max) |
| **IgG RBD** | 20 (100) | 0 (0) | --- | --- | 40 (100) | 0 (0) | --- | --- |
| **IgG nucleocapsid** | 16 (80) | 1 (5) | 3 (15) | 3.8 (0.6-8.6) | 19 (47.5) | 16 (40) | 5 (12.5) | 1.2 (0.1-6.6) |
| **IgM Spike1** | 5 (25) | 15 (75) | --- | 0.2 (0.0-4.7) | 9 (22.5) | 31 (77.5) | --- | 0.2 (0.0-4.2) |
| **IgA Spike1** | 18 (90) | 0 (0) | 2 (10) | 3.2 (0.8-5.5) | 35 (87.5) | 0 (0) | 5 (12.5) | 3.7 (0.8-10) |
| **IgM nucleocapsid** | 1 (5) | 1 (5) | 18 (90) | 0.3 (0.0-1.8) | 1 (2.5) | 36 (90) | 3 (7.5) | 0.2 (0.0-2.8) |

Positivity to the combination of IgG RBD, IgA Spike and IgM Spike was 25% at 4-9 months and 23% at 8-9 months.

The difference in IgM Spike 1 from 4-5 months to 8-9 months reached significance (p=0.009), while the difference in IgA Spike 1 between the two periods reached marginal significance (p=0.053). The differences of IgG Nucleocapsid and IgM Nucleocapsid did not reach statistical significance. The amount of IgG RBD antibody was 111.9(11.3-642.7) BAU/ml in the period 4-5 months post COVID-19 d 111.8(21.2-1820) BAU/ml in the period 8-9 months post COVID-19 (p=0.391).

The amount of RBD IgG antibodies produced at 4-5 and 8-9 months post-infection according to the degree of severity of the COVID-19 disease suffered are shown in Table 2. No statistical significance of their differences was found either within or between periods.

**Table 2.** Amount of RBD IgG Ab measured in BAU/ml as a function of disease severity, at 4-5 months and at 8-9 months post Covid-19 infection.

| COVID SEVERITY | IgG RBD positive |  | IgG (BAU/ml)<br>median (min-máx) |  | p-Value |
| --- | --- | --- | --- | --- | --- |
|  | at 4-5 months | at 8-9 months | at 4-5 months | at 8-9 months |  |
| **MODERATE n (%)** | 9 (45) | 22 (55) | 91.3 (43.9-577.9) | 130.9 (24.7-1,820) | 0.441 |
| **SEVERE n (%)** | 11 (55) | 18 (45) | 148.4 (11.3-642.7) | 94.6 (21.2-1,123.4) | 0.594 |
| **p-Value** | --- | --- | 0.503 | 0.251 | --- |

To date (February 2022) we have not detected any case of re-infection requiring a microbiological diagnosis in our study.

## Discussion

This paper presents a study of patients with a microbiological diagnosis of SARS-CoV-2 infection, who required hospital admission. A high percentage of patients had pre-existing chronic comorbidities and all of them suffered a moderate or severe course of the disease. Different studies have related the severity of the COVID-19 infection with previous the comorbidities presented. We found that hypertension and type 2 diabetes were the main diseases, coinciding with other authors such as Huang et al [8].

Half of the patients required respiratory support measures: 25% with high-flow nasal spectacles, 13% noninvasive ventilation and 13% of patients required IMV, a percentage higher than in other series [9].

After a year of follow-up, forty percent of the patients had pulmonary sequelae, Huang C et al.,[10] described that most patients (76%) are symptomatic to some degree after six months of COVID-19 infection, the most common symptomatology being fatigue and muscle weakness in 63%, insomnia in 26% and anxiety in 23%. Also, the alteration of pulmonary function presented a percentage of 22-56% of cases, being higher in those cases that had more severe cases of COVID-19 disease during the acute phase [10]. Similar results were obtained by Lombardo et al. one year after SARS-COV-2 infection in hospitalized patients, reaching a pulmonary involvement of 37% [11]. Within the pulmonary sequelae, the most frequent are interstitial thickening, ground-glass infiltrates [11] in our study we found that most frequent was mild pulmonary fibrosis.

Pulmonary fibrosis is exceptional and risk factors for its development are considered to be age, severity of COVID-19 disease, prolonged stay in ICU, need for mechanical ventilation, smoking and alcoholism [3]. In our study twenty percent of patients had mild pulmonary fibrosis and most patients suffered severe COVID-19 during the acute phase.

We have not detected any case of reinfection that required a microbiological diagnosis; nevertheless, we are aware that mild or asymptomatic self-limited cases could go unnoticed. On the other hand, authors such as Havervall et al.,[12] conclude that the vast majority of patients who present a positive Spike IgG result 8 months after COVID-19 infection have a reduced risk of both asymptomatic and mild reinfection, regardless of the severity of the initial COVID-19 infection.

Of the 40 patients who participated in the antibody analysis all presented RBD IgG after 8-9 months post COVID-19 infection, which is in line with previously published studies [13]; however, to our knowledge we are the first to quantify these antibodies in International Units (BAU/ml).

In addition, 47% of patients were positive for IgG Nucleocapsid and 2.5% for IgM Nucleocapsid, coinciding with other studies such as Ripperger et al., [14] where they report that the antibodies compared to Nucleocapsid decrease more rapidly than Ab compared to RBD. Likewise, 88% presented positivity to IgA Spike in line with other studies such as that of Dan et al., [15] which also detects it in serum in the majority of subjects 6-8 months after infection. Twenty-three percent of patients were positive for RBD IgG, Spike IgA Spike and Spike 1-RBD IgM, which could contribute to a greater capacity to neutralize the virus in these patients [16].

In the analysis of the evolution of antibodies from 4-5 months to 8-9 months we found no difference in the amount of RBD IgG detected, so that similar levels of Spike IgG A were maintained in both periods of convalescence analyzed, something already observed in other studies [15,17]. However, we did find a significant decrease in Spike 1-RBD IgM, coinciding with other authors such as Gaebler et al [18]. It has been described in the literature that during the acute phase of the disease there are higher levels of IgG to S1 and Nucleocapsid in patients with severe COVID-19 than in those with milder disease [18, 19]. However, in our study we did not find difference when the level of RBD IgG according to the severity of the COVID-19 disease suffered was compared, coinciding with other authors such as Sandberg et al.,[17] probably because all our population had a diagnosis of pneumonia and were stratified as severe or moderate COVID-19, without having patients with mild or asymptomatic disease, as well as the period after the COVID-19 infection analyzed. On the other hand we observed that the amount of RBD IgG antibodies is maintained throughout the convalescent COVID-19 phase and that the amount that remains in the medium term seems to protect against new reinfections despite of emerging viral variants.

Finally, we believe that the development or not of pulmonary fibrosis is probably independent of the amount of RBD IgG generated, since we observed that regardless of the level of antibodies maintained, a considerable proportion of COVID-19 patients developed discrete interstitial parenchymal alterations.

There are some limitations to this study. The first one is that the sample size is small, which undermines the power of the study. On the other hand, in our study the majority of patients suffered a moderate-severe degree of severity of the disease, who were those who required follow-up by the multidisciplinary consultation, so that the humoral immune response profile cannot be applied to patients with a mild degree of severity or to asymptomatic patients. Another limitation is that even if no new reinfections were detected, these may have been asymptomatic, however, these patients have a greater neutralizing capacity thanks to the positivity of IgG and IgA.

Taking these limitations into consideration, our study describes a high percentage of mild pulmonary fibrosis development in patients who presented at least one of the following conditions: persistence of pathological alterations in the chest X-ray and/or having required special ventilatory support devices during their admission.

## Data Availability

All relevant data are within the manuscript and its Supporting Information files.

## Acknowledgments

The authors would like to thank the Microbiology Laboratory Technicians (Maria Desiree Ruiz and Carmen Lorenzo), and the Infection Control nurses for their help with the procedures.

## References

1. Zhou Y, Chi J, Lv W, Wang Y. Obesity and diabetes as high-risk factors for severe coronavirus disease 2019 (Covid-19). Diabetes Metab Res Rev. 2021 Feb;37(2):e3377. doi: 10.1002/dmrr.3377. Epub 2020 Jul 20. PMID: 32588943; PMCID: PMC7361201.

2. Williamson EJ, Walker AJ, Bhaskaran K, Bacon S, Bates C, Morton CE, et al., Factors associated with COVID-19-related death using OpenSAFELY. Nature. 2020 Aug;584(7821):430–436. doi: 10.1038/s41586-020-2521-4. Epub 2020 Jul 8. PMID: 32640463; PMCID: PMC7611074.

3. Chérrez-Ojeda I, Gochicoa-Rangel L, Salles-Rojas A, Mautong H. Seguimiento de los pacientes después de neumonía por COVID-19. Secuelas pulmonares [Follow-up of patients after COVID-19 pneumonia. Pulmonary sequelae]. Rev Alerg Mex. 2020 Oct-Dec;67(4):350-369. Spanish. doi: 10.29262/ram.v67i4.847. PMID: 33631903.

4. Chandrashekar A, Liu J, Martinot AJ, McMahan K, Mercado NB, Peter L, et al., SARS-CoV-2 infection protects against rechallenge in rhesus macaques. Science. 2020 Aug 14;369(6505):812–817. doi: 10.1126/science.abc4776. Epub 2020 May 20. PMID: 32434946; PMCID: PMC7243369.

5. Maor Y, Cohen D, Paran N, Israely T, Ezra V, Axelrod O, et al., Compassionate use of convalescent plasma for treatment of moderate and severe pneumonia in COVID-19 patients and association with IgG antibody levels in donated plasma. EClinicalMedicine. 2020 Sep;26:100525. doi: 10.1016/j.eclinm.2020.100525. Epub 2020 Sep 9. PMID: 32923991; PMCID: PMC7480446.

6. Rasheed AM, Fatak DF, Hashim HA, Maulood MF, Kabah KK, Almusawi YA, et al., The therapeutic potential of convalescent plasma therapy on treating critically-ill COVID-19 patients residing in respiratory care units in hospitals in Baghdad, Iraq. Infez Med. 2020 Sep 1;28(3):357-366. PMID: 32920571.

7. WHO/BS.2020.2403 Establishment of the WHO International Standard and Reference Panel for anti-SARS-CoV-2 antibody. Available from: https://www.who.int/publications/m/item/WHO-BS-2020.2403.

8. World Health Organization. 2020. Clinical Management of COVID-19 Patients-Interim Guidance. World Health Organization, Geneva, Switzerland.

9. Huang C, Wang Y, Li X, Ren L, Zhao J, Hu Y, Zhang L, et al., Clinical features of patients infected with 2019 novel coronavirus in Wuhan, China. Lancet. 2020 Feb 15;395(10223):497–506. doi: 10.1016/S0140-6736(20)30183-5. Epub 2020 Jan 24. Erratum in: Lancet. 2020 Jan 30;: PMID: 31986264; PMCID: PMC7159299.

10. Huang C, Huang L, Wang Y, Li X, Ren L, Gu X, et al., 6-month consequences of COVID-19 in patients discharged from hospital: a cohort study. Lancet. 2021 Jan 16;397(10270):220–232. doi: 10.1016/S0140-6736(20)32656-8. Epub 2021 Jan 8. PMID: 33428867; PMCID: PMC7833295.

11. Lombardo MDM, Foppiani A, Peretti GM, Mangiavini L, Battezzati A, Bertoli S, et al., Long-Term Coronavirus Disease 2019 Complications in Inpatients and Outpatients: A One-Year Follow-up Cohort Study. Open Forum Infect Dis. 2021 Jul 16;8(8):ofab384. doi: 10.1093/ofid/ofab384. PMID: 34386546; PMCID: PMC8344801.

12. Havervall S, Ng H, Jernbom Falk A, Greilert-Norin N, Månberg A, Marking U, Laurén I, et al., Robust humoral and cellular immune responses and low risk for reinfection at least 8 months following asymptomatic to mild COVID-19. J Intern Med. 2022 Jan;291(1):72–80. doi: 10.1111/joim.13387. Epub 2021 Sep 27. PMID: 34459525; PMCID: PMC8661920.

13. Masiá M, Fernández-González M, Telenti G, Agulló V, García JA, Padilla S, et al., Durable antibody response one year after hospitalization for COVID-19: A longitudinal cohort study. J Autoimmun. 2021 Sep;123:102703. doi: 10.1016/j.jaut.2021.102703. Epub 2021 Jul 20. PMID: 34303083; PMCID: PMC8289631.

14. Ripperger TJ, Uhrlaub JL, Watanabe M, Wong R, Castaneda Y, Pizzato HA, et al., Orthogonal SARS-CoV-2 Serological Assays Enable Surveillance of Low-Prevalence Communities and Reveal Durable Humoral Immunity. Immunity. 2020 Nov 17;53(5):925-933.e4. doi: 10.1016/j.immuni.2020.10.004. Epub 2020 Oct 14. PMID: 33129373; PMCID: PMC7554472.

15. Dan JM, Mateus J, Kato Y, Hastie KM, Yu ED, Faliti CE, et al., Immunological memory to SARS-CoV-2 assessed for up to 8 months after infection. Science. 2021 Feb 5;371(6529):eabf4063. doi: 10.1126/science.abf4063. Epub 2021 Jan 6. PMID: 33408181; PMCID: PMC7919858.

16. Noval MG, Kaczmarek ME, Koide A, Rodriguez-Rodriguez BA, Louie P, Tada T,et al, Antibody isotype diversity against SARS-CoV-2 is associated with differential serum neutralization capacities. Sci Rep. 2021 Mar 10;11(1):5538. doi: 10.1038/s41598-021-84913-3. PMID: 33692390; PMCID: PMC7946906.

17. Sandberg JT, Varnaitė R, Christ W, Chen P, Muvva JR, Maleki KT, et al., SARS-CoV-2-specific humoral and cellular immunity persists through 9 months irrespective of COVID-19 severity at hospitalisation. Clin Transl Immunology. 2021 Jul 5;10(7):e1306. doi: 10.1002/cti2.1306. PMID: 34257967; PMCID: PMC8256672.

18. Gaebler C, Wang Z, Lorenzi JCC, Muecksch F, Finkin S, Tokuyama M, et al., Evolution of antibody immunity to SARS-CoV-2. Nature. 2021 Mar;591(7851):639–644. doi: 10.1038/s41586-021-03207-w. Epub 2021 Jan 18. PMID: 33461210; PMCID: PMC8221082.

19. Horton DB, Barrett ES, Roy J, Gennaro ML, Andrews T, Greenberg P, et al., Determinants and Dynamics of SARS-CoV-2 Infection in a Diverse Population: 6-Month Evaluation of a Prospective Cohort Study. J Infect Dis. 2021 Oct 28;224(8):1345–1356. doi: 10.1093/infdis/jiab411. PMID: 34387310; PMCID: PMC8436370.

